# Impact of personalized-dose vaccination in Covid-19 with a limited vaccine supply in a 100 day period in the U.S.A.

**DOI:** 10.1101/2021.01.30.21250834

**Authors:** Patrick Hunziker

## Abstract

**Background:** We aimed at minimizing loss of lives in the Covid-19 pandemic in the USA by identifying optimal vaccination strategies during a 100-day period with limited vaccine supplies. While lethality is highest in the elderly, transmission and case numbers are highest in the younger. A strategy of first vaccinating the elderly is widely used, thought to protect the vulnerable, elderly best. Despite lower immunogenicity in the elderly, mRNA vaccines retain high efficacy, implying that in the younger, reduced vaccine doses might suffice, thereby increasing vaccination counts with a given vaccine supply.

**Methods:** Using published immunogenicity data of the Moderna mRNA-1273 vaccine, we examined the value of personalized-dose vaccination strategies, using a modeling approach incorporating age-related vaccine immunogenicity, social contact patterns, population structure, Covid-19 case and death rates in the USA in late January 2021. An increase if the number of persons that can be vaccinated and a potential reduction of the individual protective efficacy was accounted for.

**Results:** Age-personalized dosing strategies reduced cases faster, shortening the pandemic, reducing the delay to reaching <100’000 cases/day from 64 to 30 days and avoiding 25’000 deaths within 100 days in the USA. In an “elderly first” vaccination strategy, mortality is higher even in the elderly. Findings were robust with transmission blocking efficacies of reduced dose vaccination between 30% to 90%, and with a vaccine supply from 1 to 3 million full dose vaccinations per day.

**Conclusion:** Rapid reduction of Covid-19 case and death rate in the USA in 100 days with a limited vaccine supply is best achieved when personalized, age-tailored dosing for highly effective vaccines is used, according to this vaccination strategy model parameterized to U.S. demographics, Covid-19 transmission and vaccine characteristics. Protecting the vulnerable is most effectively achieved by personalized-dose vaccination of all population segments, while an “elderly first” approach costs more lives, even in the elderly.

## Introduction

Vaccines against SARS-Cov2 have been developed at warp speed, and mRNA vaccines like the Pfizer BNT162b2^1^(Tozinameran) and the Moderna mRNA-1273^2^ have shown strong immunogenicity, safety and efficacy against disease. Emerging data also show protection against infection (and thus, transmission) by the immune response reached in natural infection^3^ and, presumably, vaccination^4,5^. With global case numbers and deaths reaching new peaks in January 2021, manufacturing lines are not currently capable to cover the hµge global demand, calling for optimally effective strategies for vaccine deployment. While phase I-II data have confirmed that elderly people exhibit reduced immune responses to vaccination^6^, the vaccines have been dose-optimized to achieve excellent immunity even in the elderly, sµggesting that for the younger (i.e., those ≤64 years), the standard vaccine dose may be higher than needed, as antibody levels are known to correlate with protective efficacy^7^. To achieve proof of efficacy quickly, pivotal trials were performed at “one fits all” dose levels. In the younger, sufficient immunity might be reached with lower vaccine doses^8^. Faced with the currently severe vaccine availability bottleneck, a lower dose per vaccination would translate into larger numbers of people receiving the vaccine early. As the younger, due to their social contact patterns^9,10^, drive the pandemic to a large degree, vaccinating them early while continuing to protect the vulnerable may prove to be a game changing strategy for stopping the pandemic rapidly.

Specifically, Moderna vaccine development has explored doses of 25, 50, 100 and 250µg. 25µg in the younger achieves immunity levels comparable to those seen in reconvalescent plasma in natural infection, with the latter being protected to 83% for at least 5 months^11^. 100 µg achieves high immunity levels in all age groups, even in the elderly. The pivotal study populations were protected against infection at least four months despite some reduction of the measured immunity parameter in the elderly. Notably, in the younger, a 25µg dose of the Moderna vaccine elicited an immune response level at day 57 similarly to those in patients older than 71 years at day 119 (Table), reaching >86% protection. Antibody titers after vaccination are correlated with protection from reinfection^12^. We therefore tested the hypothesis that personalized, age-personalized vaccine dosing will allow early vaccination of significantly more persons and translate into a reduction of case load, deaths and pandemic duration.

## Methods

The model (figure 1) is inspired by the SEIR (susceptible, exposed, infective, recovered) approach but has discrete structure^13,14^ with daily assessment for 100 consecutive days. It includes two age strata that are differentially parameterized for age-specific differences in social interaction, case fatality rate, and vaccine efficacy, including the interaction of the two groups through risk contact propensity.

**Legend Figure 1:**
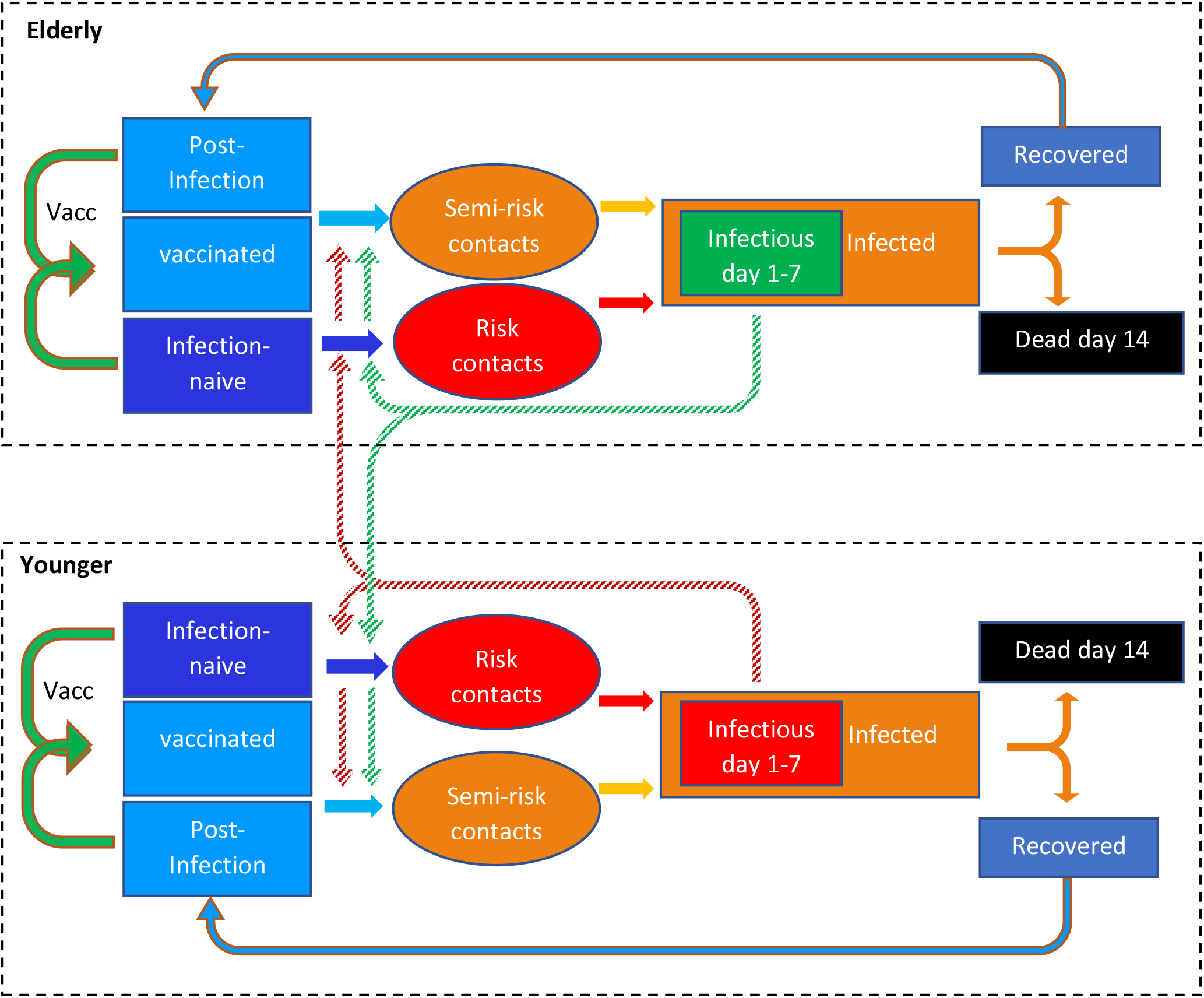
Time-discrete model inspired by the SEIR (susceptible-exposed-infective-recovered) approach, using two age strata that are parameterized differentially for vaccination protection, social contacts, and case fatality rates. Case rates are used, not infection rates, to render the approach parameterizable with available real-world data. Solid arrows indicate transition of patients to a different group; dotted arrows indicate risk contacts within and between groups.

The model was parameterized based on Moderna vaccine publications on phase I^15,16,17,18,19^ II and III^20^ studies. The model was initialized using a population size and age structure of the U.S.A with a population of 332’599’000, split into a cohort of 54’303’000, “old” persons > 64 years and of 278’296’000 “younger” persons ≤64, according to U.S. government data^21^. Covid-19 case numbers were from the U.S. Center for Disease Control^22^, the Johns Hopkins University CSSE dataset^23^ and the Oxford university “our world in data” repository^24^ and were used to initialize the model to 193’717 cases per day as per January 20, 2021. Hospitalization numbers are from the Covid Tracking project^25^ and from government sources^26^. New cases are infectious from day 1 to day 7. Stock for 1 million standard dose vaccinations per day are available. Protection by vaccination occurs from day 10.

In one analysis, protective efficacy against infection transmission of the 100µg vaccine dose was set to 95·6% in the younger and 86·2% in the elderly as published for protection against disease, and vaccine efficacy of a 25µg dose in the younger was set to 86·2% based on the levels of immunogenicity achieved in the younger compared to the immune response in the elderly vaccinated with 100µg as shown in the table. In a further analysis, protective efficacy against virus transmission after vaccination^27^ and natural infection was varied from 30% to 90% for the younger when using the 25µg dose. Based on known differences in social contacts between age groups^28^, younger persons were set to have 80% of their social contacts with the “younger” and 20% with the “old”, while for the old, contacts to other elderly and the younger were each set to be 50% each. Using risk contacts propensities at study start (derived from the numbers of non-immune, of infectious, of new cases, in each age segment) as reference, we used daily contact propensities to compute transmissions in each age group: encounters of non-immune with infectious persons were “risk contacts”, while encounters of immune with infectious persons were “semi-risk contacts”, weighting infection risk according to the protection afforded by the vaccine in that age segment. Deaths on a given day were computed from the daily case count 14 days before, using the case fatality rate in the U.S. in January 2021, approximately 1·6%, according to the relatively stable case and death counts in this period. The age-dependent Covid-19 death distribution was derived from the Center for Disease Prevention and Control data^29^, indicating a case fatality rate of 0.35% for the younger and 8.9% for the elderly, as supported by others^30^. The following scenarios were tested:

“elderly first”: starting with regular dose vaccination until 80% of the elderly are covered, then vaccinating the younger at regular dose.

“younger first”: starting with regular dose vaccination in the younger, leaving the elderly aside during the first 100 days

“personalized-dose”: in parallel, using half of the stock for each, vaccinate the elderly at full dose and the younger at quarter dose

“personalized-dose, the younger first”: starting with quarter dose vaccination in the younger, leaving the elderly aside until 80% of the younger are vaccinated, then vaccinating the elderly at full dose.

Further details of the methodology are found in the supplemental materials.

Ethics: The relevant Ethisches Kommittee Nordwestschweiz EKNW declared that computer modeling studies do not fall under the jurisdiction of Ethical Committees.

## Results

Baseline results in the “elderly first” strategy, using standard vaccine dosing, predict a cumulative death count of 153’000 over 100 days. Case numbers fall below 100’000/day on day 64 and the daily deaths fall below 1’000/day on day 55, as shown in Figure 2. In the elderly, death rates initially fall fastest compared to other scenarios, but later in the pandemic, the significant exposure of the elderly to the many infectious younger persons in the not yet vaccinated, younger, socially active population segments, and taking into account that the protective efficacy of the vaccine in the elderly is less than 100%, may lead to continued morbidity and mortality in the elderly compared to other scenarios.

**Legend Figure 2:**
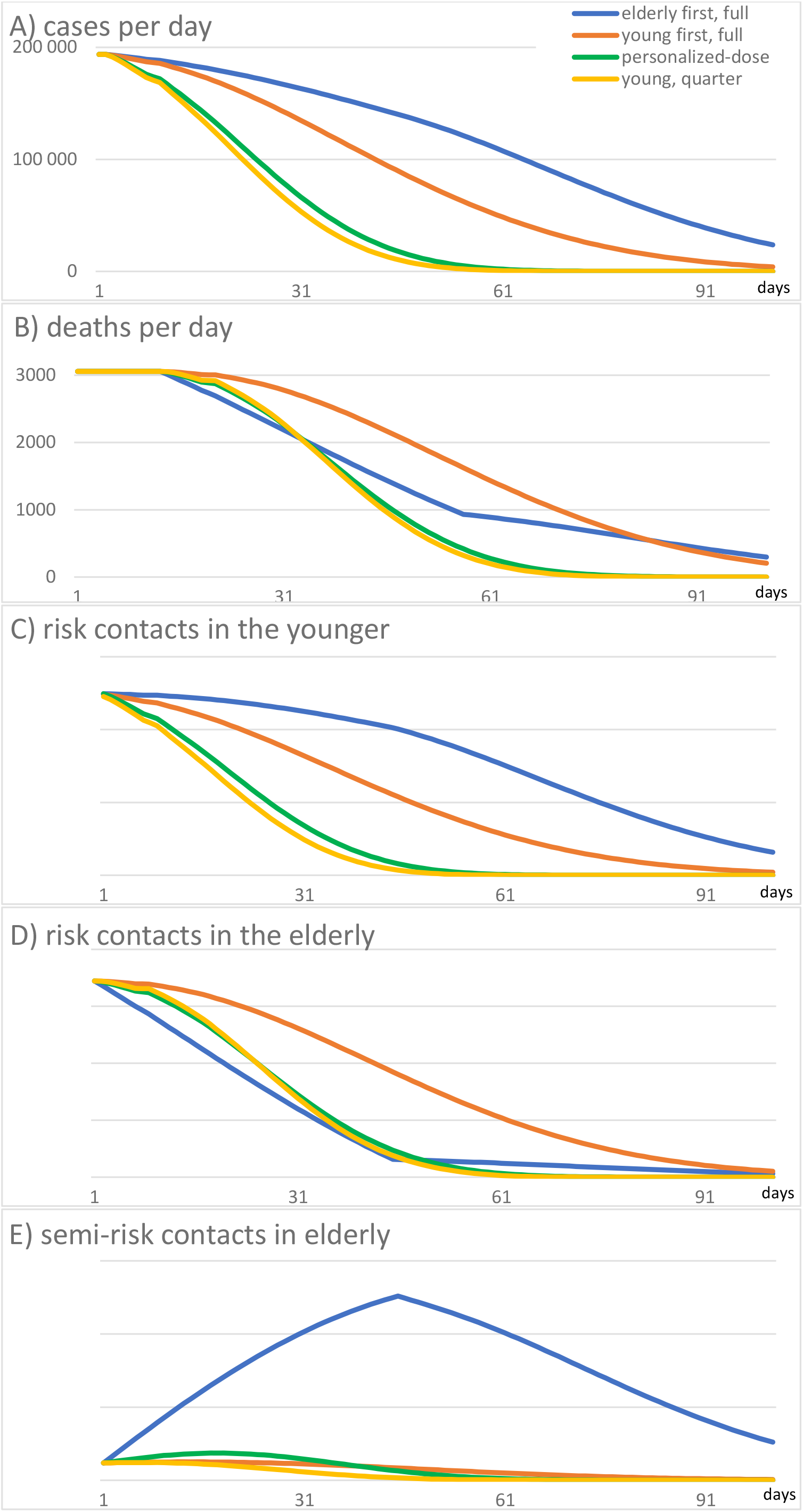
Covid-19 case numbers and deaths, and “risk contact” propensity during 100 days of a vaccination campaign modeled for the U.S.A. “elderly first” uses full dose vaccination in the elderly until 80% of the elderly are vaccinated, then switching to the younger at full dose. “younger first” vaccinates only the younger, at full dose. “personalized-dose” uses full dose in the elderly and in parallel, using quarter dose in the younger. “younger quarter” only vaccinates the younger, at quarter dose, until 80% of the younger are vaccinated. Panel A: Covid-19 cases per day, comparing the different scenarios. Note that using a reduced (quarter) dose vaccine reaches a larger proportion of the population faster, and despite a reduced efficacy per individual person, the societal impact, measured as case number reduction, is strongest for an personalized-dose, reduced dose vaccination strategy, and for the strategy with reduced dose vaccination starting solely in the younger. Panel B: Death number reduction is initially fastest by an “elderly first”, and to some degree by the “personalized-dose” strategy, while later on, the broadest approach, namely vaccinating primarily the younger, has the largest impact because it stops the pandemic most quickly. Panel C, D, E: Propensities of risk contacts (i.e., a non-immune meets an infectious person) and “semi-risk contacts” (i.e., a vaccinated or previously infected meets an infective person, acknowledging that the protection provided is less than 100%) compared to day 1. As in the strategy “elderly first”, unchecked virus propagation in the younger occurs, there is a rapid decay of “risk contacts” within the elderly, but the immunized elderly will encounter large numbers of infected younger persons, maintaining a residual risk. In contrast, the strategies that include the younger from the beginning reduce “risk” and “semi-risk” encounters significantly, in particular if the quarter dose vaccination is part of the strategy.

In the “younger first” scenario, at standard dose vaccine, case numbers fall faster in the younger, but at the expense of higher mortality in the unprotected elderly cohort throughout most of the study period, yielding a higher overall death count of 184’000, with case rates falling below 100’000/day on day 42 and death rates falling below 1000/day on day 70.

In an “personalized-dose” strategy, including reduced-dose vaccination in the young and full dose vaccination in the elderly, case numbers fall faster than with either prior strategy, and while mortality in the elderly is initially slightly higher than in the “elderly first” strategy, mortality, even in the elderly, quickly falls below the one observed in the other strategies, resulting in markedly lower cumulative deaths of 128’000 in 100 days. The milestones of <100’000 deaths/day are reached on day 30, and of <1000 deaths/day on day 49, significantly faster than in the prior scenarios. This improvement is due to the strong reduction in risk contacts, as well as the “semi-risk” contacts (a partially immune meets an infectious person) in the younger as well as in the elderly, compared to the prior scenarios, as shown in Figure 1 panels C-E.

Limiting the vaccine campaign to the younger and vaccinating them at quarter dose leads to an even faster reduction of risk contacts and case numbers, but with a similar cumulative number of deaths: 121’000 in 100 days, achieving <100’000 cases/day in 22 days and <1000 death/day in 45 days. However, this approach, as effective as it is, might convey a sense of injustice.

Sensitivity of the results were tested subsequently for a varying vaccination effectiveness parameter because information on protective effects of the vaccines against virus transmission are still sparse: assuming a protective efficacy against infection (and transmission) of quarter dose vaccination and of natural infection of only 30%, while full dose vaccination yielded infection protection levels equal to the published disease protection levels, the personalized-dose strategy yielded similar cumulative death numbers (162’000 vs 163’000) and a shortened time to <100’000 cases (50 vs 82 days) compared to the “elderly first” strategy, as shown in figure 3A. At protection levels of quarter dose vaccination against infection above 30%, the personalized-dose strategy was preferable to the “elderly first” strategy.

**Legend Figure 3:**
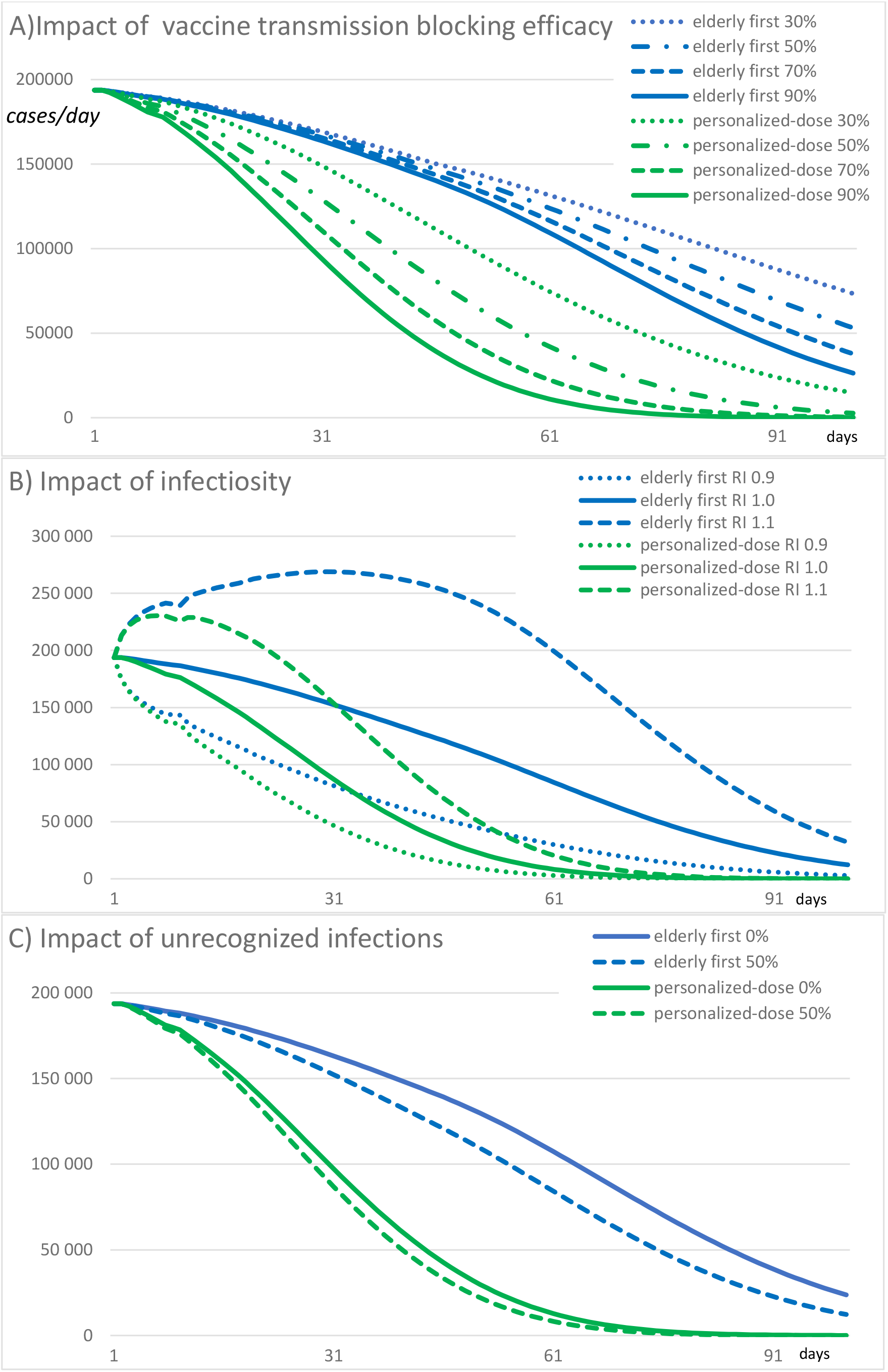
Panel A: Impact of transmission blocking effect of the vaccine in the younger on the Covid-19 cases/day arising with implementation of different vaccination scenarios in the USA. As stopping transmission in the younger is the key mechanism of success for the personalized-dose strategy, a decreasing efficacy on transmission blocking in the younger was modeled, while transmission blocking in the elderly was maintained at 86.4%, a bias in favor of the “elderly first” strategy. Note that even at low transmission blocking efficacy down to 30%, the personalized-dose strategy has a substantial impact on case numbers when compared to the “elderly first” strategies. Multiplying early vaccine recipients in the younger, i. e. choice of the vaccination strategy, predominates over transmission protection reduction in an individual person. Labels indicate the transmission blocking efficacy in the younger for a given strategy in percent. RI is relative infectiosity of the virus, compared to the baseline, wild type. Panel B: Impact of virus infectiosity on success of vaccination campaign, comparing relative infectiosities of 1.0 (corresponding to an initial reproduction number of 1.0), with viruses of increased (relative infectiosity RI=1.1) or decreased (RI=0.9) infectiosity. In viruses with higher infectiosity, differences in vaccination strategy success are amplified. Panel C: Impact of the fraction of nondiagnosed infection (FNI) in a population (adding to the number of persons with naturally acquired immunity). Varying the number of undiagnosed infections from 0 to 50% (half of infections are not confirmed as “cases”) only has a minor impact on strategy outcome.

Findings were robust when infectiosity of the virus was varied (Figure 3B) and when non-diagnosed infections were taken into account (from 0 to 1:1 with confirmed cases, figure 3C).

Results are consistent across a vaccine supply range range from 1 to 3 million full dose vaccinations per day, as shown in figure 4, showing a faster pandemic resolution with the personalized dose approach. With 3 million full vaccine doses available per day, cumulative deaths with the “elderly first” strategy are 95’000, with the “younger first” strategy, 128’000, and with the personalized-dose strategy, 83’000.

**Legend Figure 4:**
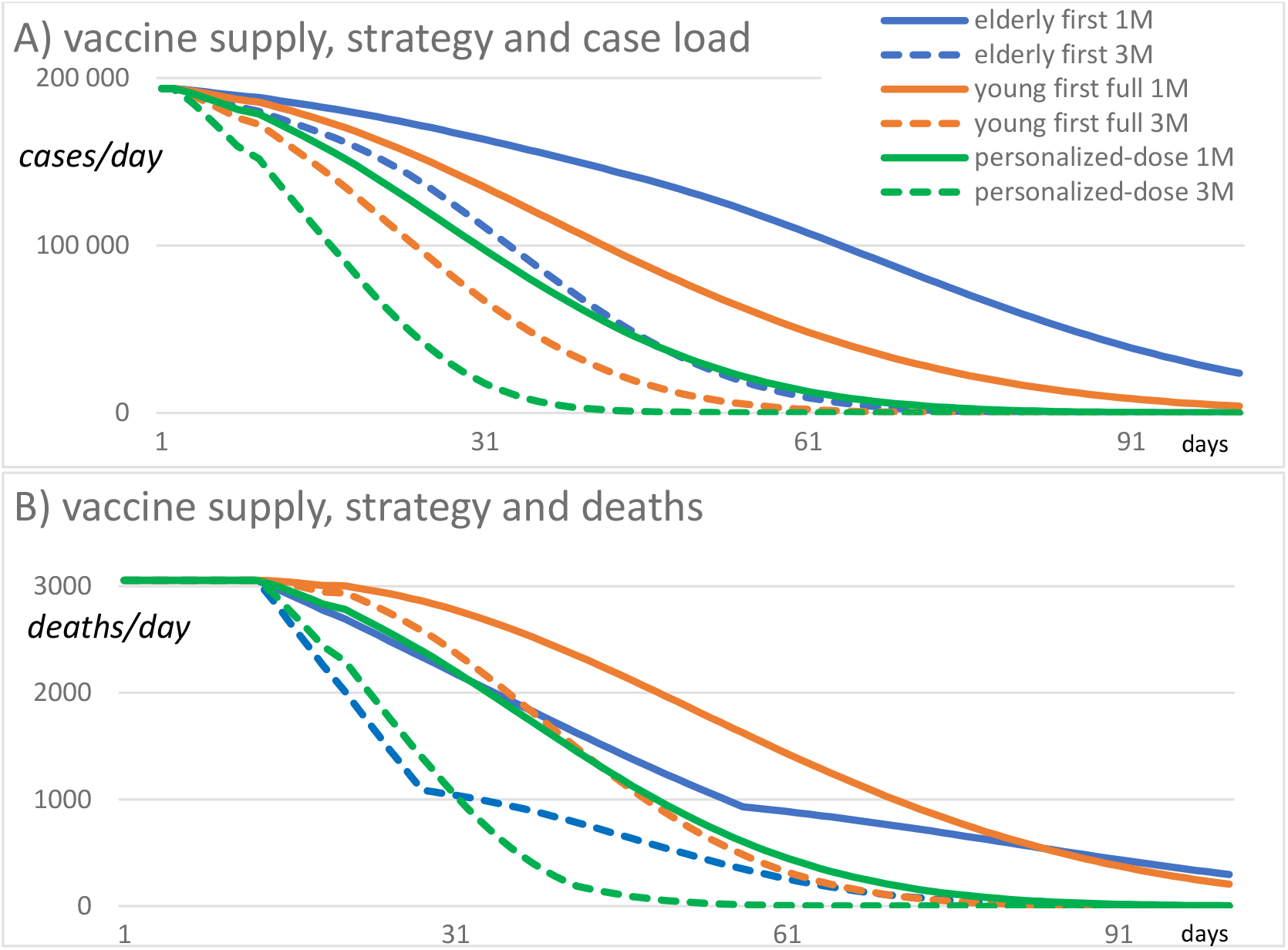
Impact of vaccine supply and strategy on daily case (A) and death rates (B). Increasing the vaccine supply accelerates all strategies but retains the distinctive advantages of the personalized-dose vaccination strategy. 1M=supply for 1 million, 3M= supply for 3 million full dose vaccinations per day. “personalized-dose” is half of vaccine supply at full dose to elderly and, in parallel, half of supply at quarter dose to younger. “elderly first” uses full dose in elderly, followed by full dose in the younger; “young full” uses all vaccine supply for the younger, at full dose.

## Discussion

The Covid-19 pandemic calls for decisive action to minimize excess deaths and long-term sequelae of the disease, protect healthcare facilities, and minimize the damage to the economy. Vaccination is a cornerstone for mastering the pandemic but identifying the optimal vaccination policies is a prodigious task. Here, we use the demographics and current epidemiologic data from the United States together with age-related social interaction patterns to build a predictive country-scale model and combine it with age-dependent immunity responses observed in the early clinical studies of the Moderna mRNA vaccine, similar to separate analyses^31^ focusing on Europe. While a widespread policy is “protect the vulnerable” implemented as “vaccinate the elderly first”^32^, we find that the vulnerable are best protected by protecting society as a whole through a broad vaccination strategy implementing personalized vaccine dosing. This was achieved by exploiting the excellent immunogenic properties of the available mRNA vaccine through fractional vaccine dosing in the younger, an approach that proved preferrable even when the lower efficacy of the reduced dose against infection and transmission is factored in.

Two of the studied scenarios, namely “personalized-dose: split the vaccine stock to vaccinate the elderly at full dose and use it at quarter dose to vaccinate as many younger as possible”, as well as the scenario “Vaccinate the younger first, at quarter dose” excelled in terms of shorting the pandemic, minimizing the number of cases and of deaths. As the “justice” of the latter strategy may be difficult to convey to the public because the “vulnerable” seem to be left out (although they even benefit from the approach), the personalized-dose approach seems to be a preferrable policy.

Emerging data indicate that the vaccines do not only protect from disease but also to a significant degree from infection and transmission. As the exact numbers for the degree of protection are not yet in, we evaluated the impact of varying degrees of infection reduction by the vaccine from 30-90% and found that the strategies proposed here are valid down to an infection transmission reduction of 30% of a vaccine.

The strong efficacy of the Moderna vaccine already at moderate immune response level, evident by its effectiveness in preventing disease within at 10-14 days after the first standard dose, before full immune response is achieved, supports these findings. Also, prior infection significantly protects against reinfection^33^, despite antibody levels in reconvalescents that are lower than those reported with quarter dose vaccination^34^ and antibody titers correlate with protection against infection^35^. Emerging data from Israel indicate that the immune response elicited in the elderly, elicited by a single dose of Pfizer’s mRNA vaccine mediates a degree of protection against infection and transmission^36^, although the age and the fact of having received only a single dose in this time window imply less than optimal antibody levels. “Fractional” dose vaccination has proven beneficial in viral poverty diseases^37,38^ further supporting the findings of this study.

The social interaction patterns have a significant impact on “risk contacts” in Covid-19 and pandemic course, as known from literature^39,40,41^ and underlined by this study. Even imperfect immunity conveyed to a significant proportion of the age groups that fire the pandemic most is therefore highly desirable.

Mutant viruses are projected to represent the next challenges. In general, vaccines are considered unlikely causes of resistance development in viruses^42^. As it is probable that mutants typically arise in persons with impaired immune responses who are unable to eliminate the virus, but coronavirus infection occurring after immunization in the healthy leads to strong immune system boost^43^, using reduced-dose vaccination is unlikely to lead to more mutants; in contrast, fastest mastering of the pandemic may be the key to prevent further mutants from emerging. As the Moderna vaccine preserves activity to the prevalent variant of concern B.1.1.7^44^ we believe that the strategy delineated here is reasonable at this time. As further mutants that might not be covered by the immune response from prior infection or by the current vaccines are expected soon^45^ and may require additional shots with modified vaccines, such iterative vaccine “boosters”, again encoding a slightly modified spike protein will further enhance the protection conveyed by prior fractional dose vaccinations. Less strain on the overloaded production lines by a fractional dose approach may free some resources to produce new, optimized vaccine batches that cover variants of concern, faster.

While we primarily base this analysis on the Moderna vaccine, we note that the Pfizer Tozinomeran vaccine has a similarly flat dose-response relationship for immunogenicity^46^ in the young, in doses from 30µg down to 10µg, measured as antibody and T cell response. This suggests that reduced-dose strategies in the young as proposed here may also be considered for the Pfizer vaccine, although generalization to further vaccine types will need additional in-depth examination of vaccine-specific immune response data.

Study limitations include assumptions that stem from phase I and II studies of limited size, and the extrapolation on clinical efficacy based on comparing measured immune titers. Case rate and case fatality rate are not ideal parameters during the course of a pandemic and mortality rates are phase-shifted to case rates; however, the substantial testing rates and the stagnation at high level of case rates and mortality in December/January 2021^47^ render their use acceptable, as information on true infection rates is still sparse. Preferably, the findings of this study are scrutinized by well-designed clinical trials although such trials would also need to be performed at “warp speed”. The most straightforward way is to allocate cities within a country to the personalized-dose approach proposed and use daily cases, deaths, hospital and ICU occupancy as continuously available endpoints, permitting rapid policy adaptation when indicated. The “off-label use” character of this approach calls for acquiring suited permits for its clinical application.

## Conclusion

Personalized-dose vaccination strategies that rely on an age-personalized vaccine dosing of a highly effective mRNA vaccine, applied to all population segments, may markedly outperform standard dose regimens that are initially focused on the elderly, according to this vaccination strategy model, parameterizied for U.S. demographics, Covid epidemiology and vaccine characteristics. By multiplying the number of persons that can be vaccinated early, this approach limits society-wide transmission earlier, and shortens pandemic duration and markedly lowers case counts and death rates, even in the vulnerable elderly.

**Table.** Immune response parameters of the Moderna vaccine by dose and age. The immune response observed in convalescent plasma compared to persons< 55 years vaccinated by the Moderna vaccine with 2x 25µg, at day 57^48^, and the immune response observed at 119 days in persons >71 years^49^. S-2P is the antigen encoded by the vaccine mRNA. RBD ELISA: receptor-binding domain binding antibodies. PsVNA50 : pseudovirus neutralization assay’s 50% inhibitory dilution. PRNT80 : live-virus plaque-reduction neutralization testing assay’s 80% inhibitory dilution. Data modified from^50^.

|  | **>71y** | **<55y** | **Convalescent plasma** |
| --- | --- | --- | --- |
|  | **2x 100µg** | **2x25µg** |  |
| **S-2P ELISA** |  | 299'751 | 142'140 |
| **RBD ELISA** | 157'964 | 183'652 | 37'857 |
| **PsVNA<sub>50</sub>** | 109 | 80·7 | 109·2 |
| **PRNT<sub>80</sub>** | 165 | 339·7 (day 43) | 158·3 |

## Data Availability

Data will be available after publication in journal, or according to specific discussion with the author.

---

GATHER checklist of information that should be included in reports of global health estimates

| Item number | Checklist item |
| --- | --- |
| **Objectives and funding** |  |
| 1 | Define the indicator(s), populations (including age, sex, and geographic entities), and time period(s) for which estimates were made. OK(methods) |
| 2 | List the funding sources for the work. OK(title page) |
| **Data inputs** |  |
| *For all data inputs from multiple sources that are synthesised as part of the study:* |  |
| 3 | Describe how the data were identified and how the data were accessed: OK(methods,research in context block) |
| 4 | Specify the inclusion and exclusion criteria. Identify all ad-hoc exclusions.<br>Provide information about all included data sources and their main characteristics. (not applicable) |
| 5 | For each data source used, report reference information or contact name/institution, population represented, data collection method, year(s) of data collection, sex and age range, diagnostic criteria or measurement method, and sample size, as relevant. OK(methods,research in context block) |
| 6 | Identify and describe any categories of input data that have potentially important biases (eg, based on characteristics listed in item 5).OK(age groupe differences in figure) |
| *For data inputs that contribute to the analysis but were not synthesised as part of the study:* |  |
| 7 | Describe and give sources for any other data inputs.(not applicable) |
| *For all data inputs:* |  |
| 8 | Provide all data inputs in a file format from which data can be efficiently extracted (eg, a spreadsheet rather than a PDF), including all relevant meta-data listed in item 5. OK(data inputs referenced data repositories in extractable format)<br>For any data inputs that cannot be shared because of ethical or legal reasons, such as third-party ownership, provide a contact name or the name of the institution that retains the right to the data.(not applicable) |
| **Data analysis** |  |
| 9 | Provide a conceptual overview of the data analysis method.(OK, in methods and supplement) A diagram may be helpful. |
| 10 | Provide a detailed description of all steps of the analysis, including mathematical formulae. This description should cover, as relevant, data cleaning, data pre-processing, data adjustments and weighting of data sources, and mathematical or statistical model(s). OK(in methods and supplement) |
| 11 | Describe how candidate models were evaluated and how the final model(s) were selected. (OK, in methods, supplement and figure) |
| 12 | Provide the results of an evaluation of model performance, if done, as well as the results of any relevant sensitivity analysis. OK (in results) |
| 13 | Describe methods of calculating uncertainty of the estimates. State which sources of uncertainty were, and were not, accounted for in the uncertainty analysis. OK(uncertainty is given as confidence interval for input data) |
| 14 | State how analytical or statistical source code used to generate estimates can be accessed. OK(data sharing statement) |
| **Results and discussion** |  |
| 15 | Provide published estimates in a file format from which data can be efficiently extracted.OK(data sharing statement) |
| 16 | Report a quantitative measure of the uncertainty of the estimates (eg, uncertainty intervals).(not applicable) |
| 17 | Interpret results in light of existing evidence. If updating a previous set of estimates, describe the reasons for changes in estimates. OK(discussion section) |
| 18 | Discuss limitations of the estimates. Include a discussion of any modelling assumptions or data limitations that affect interpretation of the estimates. OK (limitation section) |

## Supplementary materials

### Immunogenicity of fractional vaccine doses

Specifically, Moderna vaccine development has explored doses of 25, 50, 100 and 250µg. 25µg in the younger achieves immunity levels comparable to those seen in reconvalescent plasma in natural infection, with the latter being protected to 83% for at least 5 months^51^. 100 µg achieves high immunity levels in all age groups, even in the elderly. The pivotal study populations were protected against infection at least four months despite some reduction of the measured immunity parameter in the elderly. Notably, in the younger, a 25µg dose of the Moderna vaccine elicited an immune response level at day 57 similarly to those in patients older than 71 years at day 119 (Supplementary Table 1), reaching >86% protection.

## Online Methods

The model (figure 1) is inspired by the SEIR (susceptible, exposed, infective, recovered) approach with discrete structure^52,53^ with daily assessment for 100 consecutive days. It includes two age strata that are differentially parameterized for age-specific differences in social interaction, case fatality rate, and vaccine efficacy, including the interaction of the two groups through risk contact propensity. The model was parameterized based on Moderna vaccine publications on phase I^54,55,56,57,58^ II and III^59^ studies. The model was initialized using a population size and age structure of the U.S.A with a population of 332’599’000, split into a cohort of 54’303’000, “old” persons > 64 years and of 278’296’000 “younger” persons ≤64, according to U.S. government data^60^. Covid-19 case numbers were from the U.S. Center for Disease Control^61^, the Johns Hopkins University CSSE dataset^62^ and the Oxford university “our world in data” repository^63^ and were used to initialize the model to 193’717 cases per day as per January 20, 2021. Hospitalization numbers are from the Covid Tracking project^64^ and from government sources^65^. New cases are infectious from day 1 to day 7. Stock for 1 or 3 million standard dose vaccinations per day are available. Protection by vaccination occurs from day 10.

In one analysis, protective efficacy against infection transmission of the 100µg vaccine dose was set to 95·6% in the younger and 86·2% in the elderly as published for protection against disease, and vaccine efficacy of a 25µg dose in the younger was set to 86·2% based on the levels of immunogenicity achieved in the younger compared to the immune response in the elderly vaccinated with 100µg as shown in the table. In a further analysis, protective efficacy against virus transmission after vaccination^66^ and natural infection was varied from 30% to 90% for the younger when using the 25µg dose. Based on known differences in social contacts between age groups^67^, younger persons were set to have 80% of their social contacts with the “younger” and 20% with the “old”, while for the old, contacts to other elderly and the younger were each set to be 50% each. Using risk contacts propensities at study start (derived from the numbers of non-immune, of infectious, of new cases, in each age segment) as reference, we used daily contact propensities to compute transmissions in each age group: encounters of non-immune with infectious persons were “risk contacts”, while encounters of immune with infectious persons were “semi-risk contacts”, weighting infection risk according to the protection afforded by the vaccine in that age segment. Deaths on a given day were computed from the daily case count 14 days before, using the case fatality rate in the U.S. in January 2021, approximately 1·6%, according to the relatively stable case and death counts in this period. The age-dependent Covid-19 death distribution was derived from the Center for Disease Prevention and Control data^68^, indicating a case fatality rate of 0.35% for the younger and 8.9% for the elderly, as supported by others^69^. The following scenarios were tested:

Robustness of the model was tested by varying the protection degree of the vaccination, the infectiosity, and the prevalence of unrecognized infection, shown in figure 3.

A decreasing efficacy on transmission blocking with reduced-dose vaccination in a range from 90% down to 30% in the younger was modeled, while transmission blocking in the elderly was maintained at 86.4%, a bias in favor of the “elderly first” strategy.

The impact of virus infectiosity on success of a vaccination campaign was analyzed by comparing relative infectiosities(RI) of 1.0 (corresponding to an initial reproduction number of 1.0), with virus strains of increased (relative infectiosity RI=1.1) or decreased (RI=0.9) infectiosity. In virus strains with higher infectiosity, differences in vaccination strategy success are amplified.

The impact of the fraction of nondiagnosed infection (FNI) in a population (that increase the number of persons with naturally acquired immunity) was assessed. Varying the number of undiagnosed infections from 0 to 50% (half of infections are not counted as confirmed “cases”) only has a minor impact on strategy outcome.

**Supplementary table 1:** Immune response parameters of the Moderna vaccine by dose and age. The immune response observed in convalescent plasma compared to persons< 55 years vaccinated by the Moderna vaccine with 2x 25µg, at day 57^70^, and the immune response observed at 119 days in persons >71 years^71^. S-2P is the antigen encoded by the vaccine mRNA. RBD ELISA: receptor-binding domain binding antibodies. PsVNA50 : pseudovirus neutralization assay’s 50% inhibitory dilution. PRNT80 : live-virus plaque-reduction neutralization testing assay’s 80% inhibitory dilution.

|  | >71y | <55y | Convalescent plasma |
| --- | --- | --- | --- |
|  | 2x 100µg | 2x25µg |  |
| **S-2P ELISA** |  | 299’751 | 142’140 |
| **RBD ELISA** | 157’964 | 183’652 | 37’857 |
| **PsVNA<sub>50</sub>** | 109 | 80·7 | 109·2 |
| **PRNT<sub>80</sub>** | 165 | 339·7 (d43) | 158·3 |

<sup>51</sup> Lumley S F, O’Donnell D, Stoesser NE, et al. Antibody status and incidence of SARS-CoV-2 infection in health care workers. *N Engl J Med* 2020; DOI: 10.1056/NEJMoa2034545
<sup>52</sup> Ibeas, A., de la Sen, M., Alonso-Quesada, S. et al. Stability analysis and observer design for discrete-time SEIR epidemic models. *Adv Differ Equ* **2015**, 122 (2015). https://doi.org/10.1186/s13662-015-0459-x
<sup>53</sup> Hu, Z, Teng, Z, Jiang, H: Stability analysis in a class of discrete SIRS epidemic models. *Nonlinear Anal., Real World Appl.* 2012; **13**, 2017–33
<sup>54</sup> Jackson L A, Anderson EJ, Rouphael NG, et al. An mRNA vaccine against SARS-CoV-2—preliminary report. *N Engl J Med* 2020; DOI: 10.1056/NEJMoa2022483.
<sup>55</sup> Anderson E J, Rouphael NG, Widge AT, al. Safety and immunogenicity of SARS-CoV-2 mRNA-1273 vaccine in older adults. *N Engl J Med N Engl J Med* 2020; **383**: 2427–38.

